# Quantifying drug-related problems in community pharmacy practice: A pharmacoepidemiological study from Greece

**DOI:** 10.64898/2026.07.28.26359112

**Authors:** Vasileios A Antoniadis, Sharon Haughey

## Abstract

**INTRODUCTION:** Drug-related-problems (DRPs) are encountered daily in prescription medicines at the community pharmacy but they are neither documented nor evaluated systematically. Population ageing in Greece is expected to increase polypharmacy and consequently DRPs, thus deteriorating patients’ health and pressuring the already underfunded health system. Community pharmacists are ideally placed to review patients’ medicines and in collaboration with physicians resolve identified issues.

**OBJECTIVES:** The primary outcomes of the study were to define the number and nature of DRPs identified in prescription medicines at an urban community pharmacy in Greece. Secondary outcomes included the assessment of severity of errors and, for the evaluation part of the project, planned interventions to address them.

**METHODS:** Our study was divided into two parts: the main part which included the evaluation of the current service and a following small-scale improvement project. For the first aspect, all prescription medicines dispensed from the pharmacy within one month were analyzed, using primary data extracted directly from pharmacy prescription and medication records. Medication characteristics as indicated in prescriptions (name of drug, dosage, formulation, dosing interval) and patients’ medication records were utilized as sources of information. The identification of DRPs was based mainly on explicit criteria and reliable sources. Potential DRPs were classified following the Pharmaceutical Care Network Europe (PCNE) classification system (2019) while their severity was assessed and documented according to a tool developed by Abdel-Qader *et al*. (2010). A comprehensive approach of medication review as defined in “Polypharmacy Guidance: Realistic Prescribing” (Scottish Government Polypharmacy Model of Care Group, 2018) and patient interviews as an additional source was followed for the 4 selected patients in the second part of the study.

**RESULTS/DISCUSSION:** In 635 prescriptions containing 1464 medicines we identified 364 DRPs in 529 different patients. The most common problem was related with a possible adverse drug event (P2.1, 62.1%) while 62% of the causes that led to problems belonged to inappropriate combination of drugs (C1.4), inappropriate drug (C1.2), drug dose too high (C3.2) and duration of treatment too long (C4.2). Most DRPs were characterized as significant (n=202, 55.5%) and minor (n=110, 30.2%) while the intervention “drug changed to” (I3.1, 25.8%) was the most frequent among the planned interventions. In the improvement project, 39 DRPs were identified in the 4 enrolled patients, including errors associated with potential inappropriate omissions and absence of monitoring not discovered in the first part of the study.

**CONCLUSION:** We quantified the potential DRPs encountered in a community pharmacy and discovered that for every 4 medicines dispensed 1 potential DRP was identified. Even if all errors will not result in harm, they will create additional work and possibly lead to prescribing cascade. Additional sources of information, like the patient interviews we incorporated in the improvement project, are expected to increase the number and consistency of findings. Finally, the value of community pharmacists as a vital component of primary care was pointed out.

## INTRODUCTION

The recognition and resolution of drug-related-problems (DRPs) is at the core of pharmaceutical care (Cipolle *et al*., 2012). The Pharmaceutical Care Network Europe (PCNE) foundation (2019) defines a DRP as “an event or circumstance involving drug therapy that actually or potentially interferes with desired health outcomes”. Adverse drug events, a component of DRPs, are a major cause of hospital admissions and it is estimated that about 15-40% of those encountered in ambulatory care are preventable (Taché *et al*., 2011). The annual cost of medication errors globally is estimated today at 42 billion US dollars (Aitken and Gorokhovich, 2012) while population ageing followed by multimorbidity and polypharmacy is expected to increase this number (Barnet *et al*., 2012).

Greece is facing a demographic problem and according to current evidence its population projections are pessimistic (Balourdos *et al*., 2019; Eurostat, 2019). Moreover, considering that government funding of public health has declined rapidly since financial recession has started in 2008 (Economou, 2017) and that mortality from adverse events during medical treatment has risen (Laliotis *et al*., 2016), the necessity of rational use of medicines is even more imperative. The World Health Organization (WHO) (2017) has launched an initiative to reduce globally medication related errors by 50% in the next 5 years (Donaldson *et al*., 2017) in which pharmacists, due to their expertise in medicines, are expected to play a vital role. A recent systematic review which evaluated the effect of pharmacists’ interventions on patient outcomes showed improvement in blood pressure control but the results were inconclusive regarding other endpoints like in glycated haemoglobin, hospital readmissions or adverse drug effects (Barra *et al*, 2018). This might be explained by the lack of homogeneity of individual studies like in the methods used to measure impact, individual patients’ characteristics or types of interventions offered.

In Greece, only 6% of physicians are general practitioners (GPs) or family doctors (OECD, 2017), thus patients have their medicines prescribed from different specialized physicians which hampers the efficient overview of their pharmacotherapy. Potential DRPs are often identified in prescription medicines at the community pharmacy at the dispensing process. However, they are neither documented nor assessed mostly due to excessive workloads and lack of training. We quantified and subsequently classified, according to PCNE classification system, the potential DRPs that we identified in prescription medicines dispensed from a community pharmacy within a single month. For the first, main, part of the study, a retrospective clinical analysis was undertaken on medicines during which patient’s medication history and prescription data were used as the only sources of information. The second aspect of the study included a small-scale improvement project and a more thorough analysis of medication regimen by employing patient interviews as an additional source of information. In both cases, the clinical appraisal was based on both implicit and explicit criteria with emphasis on the latter to increase the credibility of the study and allow dissemination of results on other settings. The project was designed to assess if a trained community pharmacist was able to detect, classify and assess DRPs.

## AIM OF THE STUDY

The primary outcomes for both parts of the study were the number and nature of identified DRPs. Secondary outcomes included the severity of errors while for the service evaluation project we also proposed interventions to resolve the identified DRPs.

## METHODS

### Context

The project was undertaken at an urban community pharmacy in Greece. The pharmacy dispenses prescription drugs both for acute and chronic conditions. However, high-cost drugs including monoclonal antibodies, antineoplastic agents and immunosuppressants are provided exclusively by public pharmacies regulated by government (SFEE, 2017). Almost all prescriptions that are filled in at the pharmacy, either one time or repeat, are prescribed by physicians through an electronic format and printed in paper. The prescriber indicates the diagnosis for each prescribed drug along with the active ingredient to be dispensed, the dose, the formulation of the drug and the relevant instructions for use. Since the vast majority of physicians in Greece are specialized (OECD, 2017), we assumed that the proximity of the community pharmacy to certain doctors’ offices might affect the type of dispensed drugs and thus the nature of identified DRPs.

### Interventions

All prescription drugs that were dispensed from the community pharmacy during April 2018 were analyzed clinically. Community pharmacists act as the last line of defense on the supply of medicines to patients and the clinical evaluation of medications by them might identify a preventable error before it reaches and potentially harm the patient. Patient (age, sex, number of prescribed drugs) and medication characteristics (name of drug, dosage, formulation, dosing interval) were collected manually from electronic prescriptions. These data along with each patient’s medication record were utilized as sources of information on which the clinical review was based on. Patients’ medication records across different community pharmacies are not linked thus we had data only for drugs dispensed by our pharmacy. The clinical evaluation was conducted by the pharmacist who holds a MPharm degree, a PG Diploma in Advanced Clinical Pharmacy Practice and three years’ experience in community pharmacy. Additional training for the identification of DRPs was not undertaken while the rest of the pharmacy personnel was not involved in the project. Prescriptions that contained vaccines were not included in our analysis since the pharmacy does not possess patients’ immunization history which hinders the identification of issues related with their use. Influenza vaccines may be bought from the pharmacy, as per patient’s request, or prescribed by a doctor so it was decided not to include them in the service evaluation. Similarly, over the counter (OTC) medicines, like ibuprofen, may be either prescribed by a doctor or purchased directly from pharmacy. In our analysis we included only those that were prescribed by physicians. Both repeat and single prescriptions were analyzed. Prescriptions that contain repeat medications generally do not include drugs for acute conditions and the same occurred in our sample of prescriptions. The separation between acute and repeat prescriptions was made by the pharmacist based on the diagnosis, as indicated on the prescription, and on patient’s medication history. Certain patients filled in their repeat prescription at the community pharmacy twice in the same month. The second time was excluded from our analysis if the drugs have been appraised before, to avoid overrepresentation of them in the final total. Moreover, certain regular patients, of whom repeat drugs were analyzed, presented at the pharmacy to fill in a prescription with acute drugs. Even though these drugs were evaluated and counted as separate in the final aggregation, these patients were not calculated as separate to avoid their double representation in the final count.

### Improvement Project

For the improvement part of the project, 4 regular patients of the pharmacy were recruited. To take part in this aspect of the study a patient should take concomitantly 4 or more repeat prescription drugs and have full capacity to give an informed consent. A 10-minute interview, guided by “Polypharmacy Guidance: Realistic Prescribing” (Scottish Government Polypharmacy Model of Care Group, 2018), took place with the selected patients at the pharmacy as part of our usual counselling session. Patient interviews were utilized as an additional source of information while patient and medication records were employed in the same way as in the evaluation part of this study. The pharmacist did not have prior experience in conducting structured medication reviews.

### Detection of DRPs

For the identification of DRPs both implicit and explicit criteria were employed, with emphasis on the latter. Implicit criteria included pharmacist’s clinical judgement whereas explicit criteria included tools, reliable sources of information and official guidelines. The latest version of the validated STOPP (Screening Tool of Older Person’s Prescriptions)/START (Screening Tool to Alert doctors to Right Treatment) criteria, and specifically those criteria that did not require access to patients’ clinical records were utilized on elderly patients (O’Mahony *et al*., 2015). The STOPP/START criteria were preferred over other relevant prescribing criteria because they have been applied on community pharmacies in Europe, which used similar methods of data collection (Ryan *et al*., 2013), while their effects were found to be superior over other tools on preventing ADRs (Hamilton *et al*., 2011). The British National Formulary (BNF), BNF for children and summary of product characteristics (SPC) were used as references to resolve issues regarding maximum or minimum recommended dosage, dosing interval or off-label use. The pharmacy’s dispensing software does not have embedded any drug-drug interaction checker thus all drugs were assessed for drug-drug interactions using the online Stockley’s Interactions Checker. The grading of severity of each interaction generated by this tool was used as guide rather than as an absolute indicator for its consequent classification and assessment of significance. When clinical issues could not be resolved using the above sources, Cochrane Library and subsequently PubMed were explored to address them. Prescription guidelines issued by NICE were used when evaluating the appropriateness of pharmacotherapy for chronic conditions like asthma and dementia while the official Greek guidelines for infectious diseases (KEELPNO, 2015) was our reference for prescribed antibiotics. Anticholinergic burden was estimated for patient’s on multiple drugs utilizing an appropriate scale (Scottish Government Polypharmacy Model of Care Group, 2018). All the explicit criteria applied in the study are available in a supplementary material (Supplement 1).

### Classification of outcomes

There is variability in the definition of DRPs followed by different studies (Basger *et al.,* 2014). The definition of drug-related-problems we adopted was the one proposed by the PCNE, according to which “a drug-related-problem is an event or circumstance involving drug therapy that actually or potentially interferes with desired health outcomes”. The PCNE classification system V8.03 was selected among other similar tools that have been also validated and applied in various settings. This system differentiates the cause, or the process that may lead to a health issue, from the problem which is the actual or the potential adverse outcome. Throughout this paper, the terms “problem” and “drug-related-problems (DRPs)” are used to define the outcome. Thus, for every DRP both the cause and the consequent interventions to resolve it were recorded. This classification system has been applied widely in community pharmacies, has a clear definition of DRPs and also incorporates distinct coding for the problem, cause and intervention section facilitating their subsequent analysis. All the available subdomains for problems were utilized in our study whereas only 18 subdomains for causes were used since the rest were considered inapplicable within the current framework. For the purpose of this study we defined the subdomains P3.3 and C8.2 as “Unnecessary increased pill burden” and “Cost-effective alternatives available”, respectively. The proposed interventions were confined to “drug level” and “other intervention” subdomains while the subdomain I4.1 was defined for this study as “Increased monitoring”. The dispensed prescription medicines and those implicated with an error were classified according to the Anatomical Therapeutic Chemical (ATC) classification system, as it is defined by the World Health Organization (WHO). Pharmaceutical products containing two or more active ingredients were classified according to ATC codes for combination products. If just one of these ingredients was implicated in a DRP, it was assigned an ATC code as a plain product. When two or more drugs were implicated in one DRP, like in drug-drug interactions, we assigned them both as contributors of the problem, but we counted this issue as one DRP in the final total.

While the quantification of DRPs allows the evaluation of the current service, the significance of encountered DRPs is also considered of high importance as it increases the clinical relevance of study findings contributing to a more robust dissemination of results (Garfield *et al.,* 2013). The significance of DRPs was evaluated by using a tool developed by Abdel-Qader *et al*. (2010) and modified to fit our study methodology. This tool was considered convenient to our project, showed high-rater reliability and does not require input from many healthcare professionals (Garfield *et al.,* 2013).

### Analysis

All the data were managed and analyzed statistically at Microsoft Excel for Office 365. For the documentation of identified DRPs and planned interventions we followed the PCNE classification system preexisting coding while for the clinical severity of errors we coded the relevant scale. The basic characteristics of the study population were described with counts, percentages, means and standard deviations while the findings were described as absolute (counts) and relative (percentages).

### Ethical Considerations

Regarding the service evaluation, the School of Pharmacy Ethics Committee of Queen’s University of Belfast reviewed the project and deemed that a full ethical review was not required. For the second part which included patient interviews, a protocol was developed and approved by the School of Pharmacy Ethics Committee (Reference no: 008PMY2019). The patients that took part in the improvement project signed an informed consent prior to their participation. Full patient anonymity was maintained since only patient’s age and sex were recorded on both cases.

## RESULTS

### Basic data

Females that filled in a prescription at the community pharmacy accounted for 58% of the evaluation project population (Table 1). Overall, 529 different patients presented a prescription at the pharmacy during the study period with a mean age of 59.8 years (range 1- 98, SD=20.6). Almost half of those patients (n=243, 46%) belonged to 41-65 age group followed by those aged between 66 and 80 years (n=127, 24%). In total, 635 prescriptions containing 1464 prescribed drugs were analyzed clinically. Of these prescriptions, 460 (72.4%) were repeat prescriptions for chronic medical conditions whereas only 175 (27.5%) of them contained drugs intended for acute diseases (Table 1). The average number of medications per patient was 2.8 (range 1-12) while most patients (57%) were prescribed with 2 or less drugs. Of the 1464 drugs dispensed, more than one third of them (34.6%) belonged to ATC- group C (cardiovascular system) followed by the drugs of group A (alimentary tract and metabolism) (16.1%) (Table 1). Considering the 2^nd^ level of ATC classification, the most frequently prescribed drugs belonged to lipid modifying agents (C10) (10.85%), agents acting on the renin-angiotensin system (C09) (10.72%) and antithrombotics (B01) (7.58%).

**Table 1.** Basic characteristics for patients included into the service evaluation (N=529)

|  | **All patients**<br>**N= 529 (%)** | **With DRP**<br>**N=222 (%)** | **Without**<br>**DRPs N=307**<br>**(%)** |
| --- | --- | --- | --- |
| Female sex | 306 (58) | 123 (55) | 183(60) |
| Prescriptions | 635 | 260 | 375 |
| Repeat | 460 (72.4) | 191 (73.5) | 269 (71.8) |
| One-time | 175 (27.5) | 69 (26.5) | 106 (28.2) |
| Prescription medicines | 1464 | 840 | 624 |
| Age (years), mean ± SD (range) | 59.8 ± 20.6<br>(1-98) | 61.9 ± 23.3<br>(2-98) | 58.4 ± 18.2<br>(1-94) |
| 0-18 | 35 (6.6) | 21 (9.4) | 14 (4.5) |
| 19-40 | 36 (6.8) | 12 (5.4) | 24 (7.8) |
| 41-65 | 243 (46) | 70 (31.5) | 173 (56.3) |
| 66-80 | 127 (24) | 66 (29.7) | 61 (19.9) |
| >80 | 88 (16.6) | 53 (23.8) | 35 (11.4) |
| Number of medications per patient |  |  |  |
| $\leq 2$ | 302 (57) | 82 (36.9) | 220 (71.7) |
| 3-6 | 193 (36.5) | 112 (50.5) | 81 (26.4) |
| 7-10 | 33 (6.2) | 27 (12.2) | 6 (2) |
| $\geq 11$ | 1 (0.2) | 1 (0.4) | - |
| Number of medical conditions per patient, mean ± SD (range) | 2±1.4 (1-8) | 2.5±1.7 (1-8) | 1.6± 1.0 (1-6) |
| Number of medicines per patient, mean ± SD (range) | 2.8±2 (1-12) | 3.8±2.2 (1-12) | 2±1.4 (1-8) |
| Prescribed medicines according to ATC main groups |  |  |  |
| A: Alimentary Tract and Metabolism | 236 (16.1) | 138 (16.4) | 98 (15.7) |
| B Blood and blood forming organs | 132 (9) | 69 (8.2) | 63 (10.1) |
| C: Cardiovascular System | 506 (34.6) | 269 (32) | 237 (38) |
| D: Dermatologicals | 24 (1.6) | 7 (0.8) | 17 (2.7) |
| G: Genito urinary system and sex hormones | 43 (2.9) | 20 (2.4) | 23 (3.7) |

**Table 1** (continued)
|  | **All patients**<br>**N= 529 (%)** | **With DRP**<br>**N=222 (%)** | **Without**<br>**DRPs N=307**<br>**(%)** |
| --- | --- | --- | --- |
| H: Systemic hormonal preparations,<br>excluding sex hormones and insulins | 36 (2.5) | 21 (2.5) | 15 (2.4) |
| J: Anti-infective for systemic use | 92 (6.3) | 56 (6.7) | 36 (5.8) |
| L: Antineoplastic and<br>immunomodulating agents | 10 (0.7) | 1 (0.1) | 9 (1.4) |
| M: Musculo-skeletal system | 66 (4.5) | 27 (3.2) | 39 (6.2) |
| N: Nervous system | 170 (11.6) | 126 (15) | 44 (7) |
| P: Antiparasitic products, insecticides<br>and repellents | 2 (0.1) | 2 (0.2) | - |
| R: Respiratory system | 131 (8.9) | 94 (11.2) | 37 (5.9) |
| S: Sensory organs | 16 (1.1) | 10 (1.2) | 6 (1) |
| V: Various | - | - | - |

### DRPs

We discovered 364 DRPs in 529 patients (mean number of DRPs per patient=0.69) (Table 1). The 364 DRPs were identified among 222 patients of the 529 patients included in the evaluation part, ending up in 1.64 DRPs per patient (Table 1). In 307 patients (58% of total patients), no DRPs were identified whereas in 95, representing the 18% of all patients, we identified two or more DRPs (Table 4). In total, 456 medicines were involved in the 364 DRPs while the unique medicines, as indicated by an ATC coding, that caused a problem were 170. This is explained by the fact that when two or more medicines were involved in a DRP, for example in drug-drug interactions or duplication of same active ingredient, we assigned them both as contributors to the DRP. Among the unique medicines, 42 belonged to the nervous system, followed by those of the cardiovascular system (n=31). The medicines most frequently implicated with a DRP was bromazepam (n=19) followed by esomeprazole (n=18) and hydrochlorothiazide (n=14) (Table 4). For every potential DRP, just one cause has been assigned to it, so 364 causes have been documented in total (Table 2).

**Table 2.**
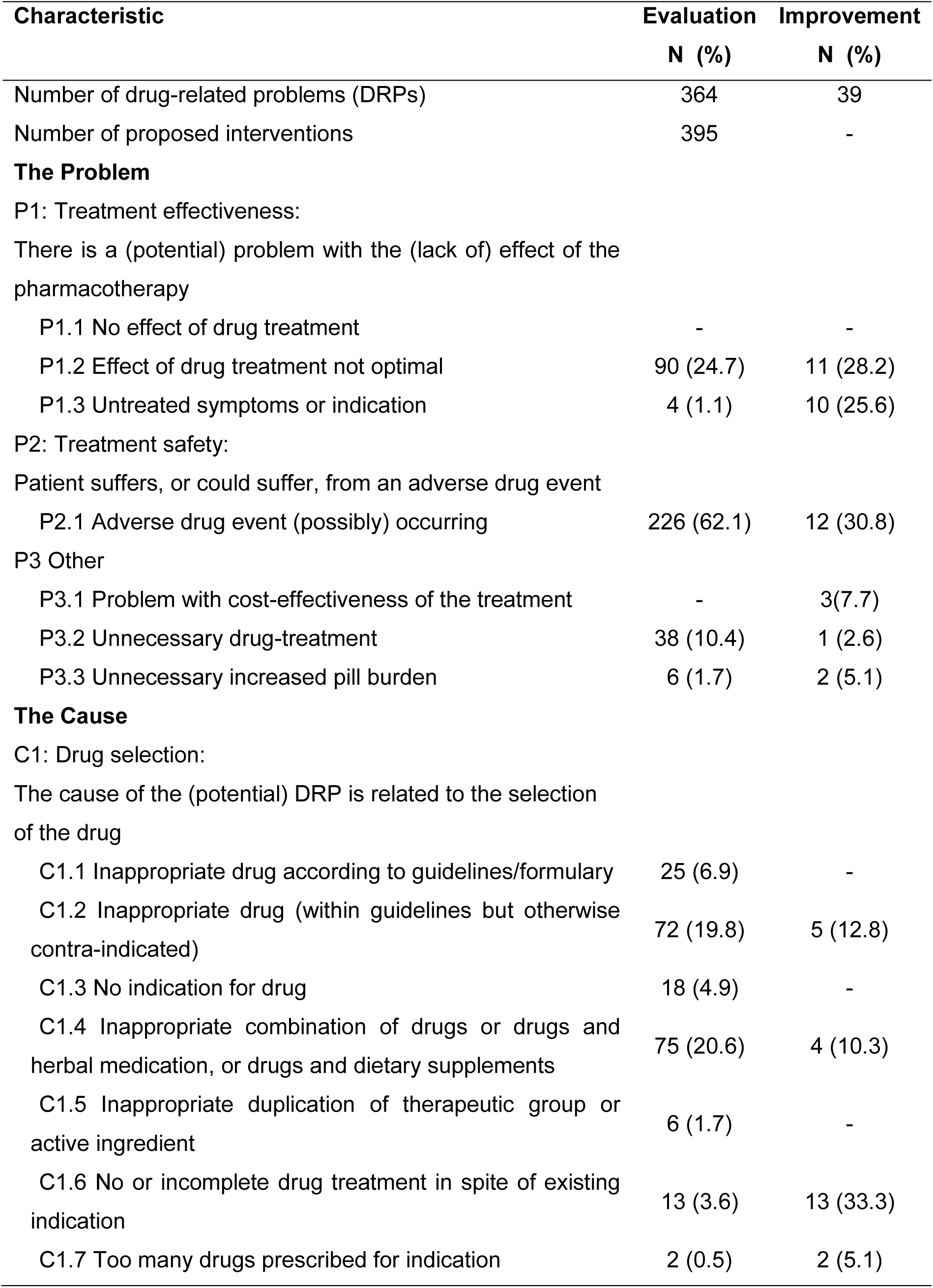
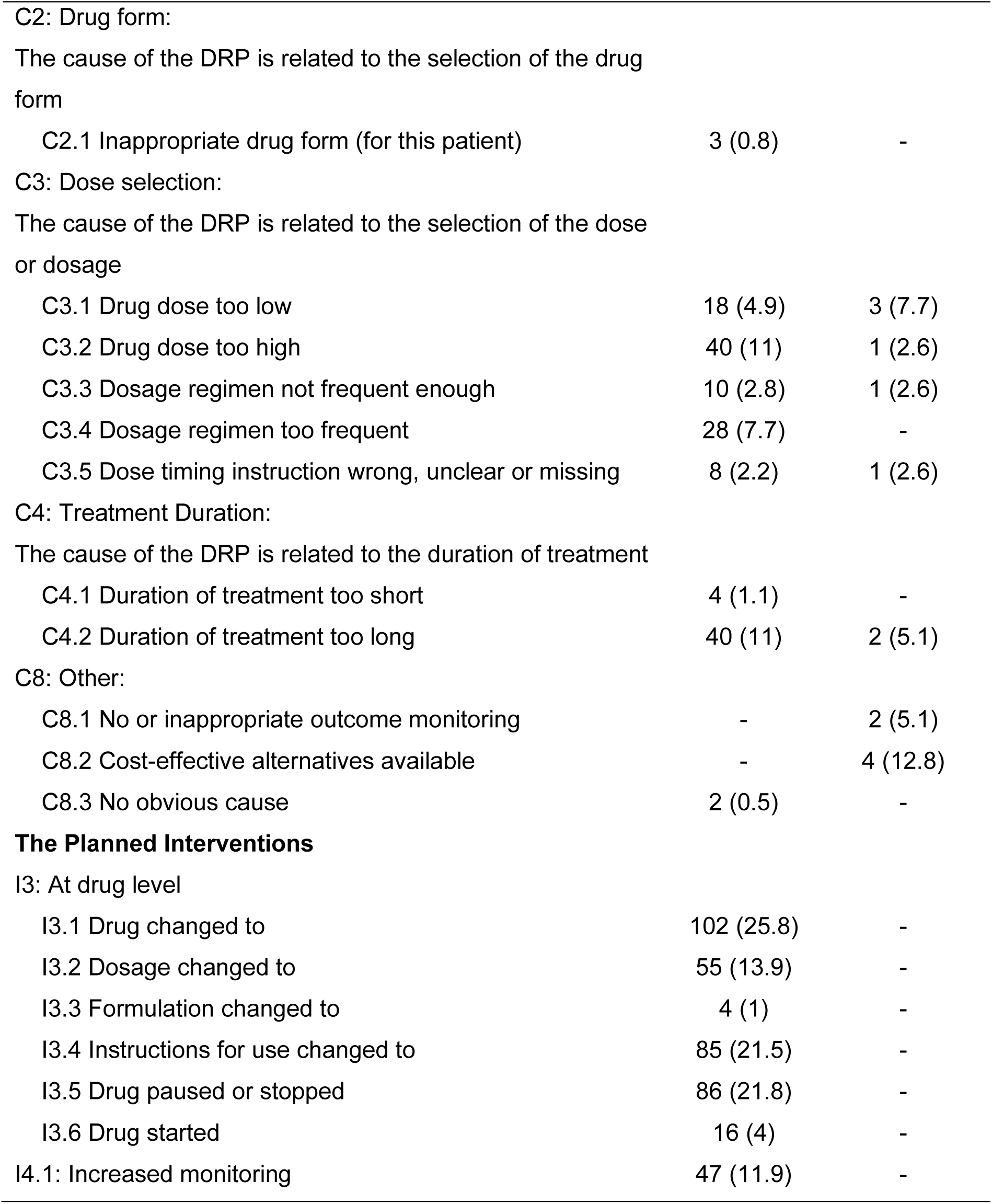
Classification of potential DRPs according to PCNE V8.03.

| Characteristic | Evaluation | Improvement |
| --- | --- | --- |
|  | N (%) | N (%) |
| Number of drug-related problems (DRPs) | 364 | 39 |
| Number of proposed interventions | 395 | - |
| **The Problem** |  |  |
| P1: Treatment effectiveness: |  |  |
| There is a (potential) problem with the (lack of) effect of the pharmacotherapy |  |  |
| P1.1 No effect of drug treatment | - | - |
| P1.2 Effect of drug treatment not optimal | 90 (24.7) | 11 (28.2) |
| P1.3 Untreated symptoms or indication | 4 (1.1) | 10 (25.6) |
| P2: Treatment safety: |  |  |
| Patient suffers, or could suffer, from an adverse drug event |  |  |
| P2.1 Adverse drug event (possibly) occurring | 226 (62.1) | 12 (30.8) |
| P3 Other |  |  |
| P3.1 Problem with cost-effectiveness of the treatment | - | 3(7.7) |
| P3.2 Unnecessary drug-treatment | 38 (10.4) | 1 (2.6) |
| P3.3 Unnecessary increased pill burden | 6 (1.7) | 2 (5.1) |
| **The Cause** |  |  |
| C1: Drug selection: |  |  |
| The cause of the (potential) DRP is related to the selection of the drug |  |  |
| C1.1 Inappropriate drug according to guidelines/formulary | 25 (6.9) | - |
| C1.2 Inappropriate drug (within guidelines but otherwise contra-indicated) | 72 (19.8) | 5 (12.8) |
| C1.3 No indication for drug | 18 (4.9) | - |
| C1.4 Inappropriate combination of drugs or drugs and herbal medication, or drugs and dietary supplements | 75 (20.6) | 4 (10.3) |
| C1.5 Inappropriate duplication of therapeutic group or active ingredient | 6 (1.7) | - |
| C1.6 No or incomplete drug treatment in spite of existing indication | 13 (3.6) | 13 (33.3) |
| C1.7 Too many drugs prescribed for indication | 2 (0.5) | 2 (5.1) |

**Table 2 (Continued)**
| **Characteristic** | **Evaluation** | **Improvement** |
| --- | --- | --- |
|  | **N (%)** | **N (%)** |
| C2: Drug form: |  |  |
| The cause of the DRP is related to the selection of the drug form |  |  |
| C2.1 Inappropriate drug form (for this patient) | 3 (0.8) | - |
| C3: Dose selection: |  |  |
| The cause of the DRP is related to the selection of the dose or dosage |  |  |
| C3.1 Drug dose too low | 18 (4.9) | 3 (7.7) |
| C3.2 Drug dose too high | 40 (11) | 1 (2.6) |
| C3.3 Dosage regimen not frequent enough | 10 (2.8) | 1 (2.6) |
| C3.4 Dosage regimen too frequent | 28 (7.7) | - |
| C3.5 Dose timing instruction wrong, unclear or missing | 8 (2.2) | 1 (2.6) |
| C4: Treatment Duration: |  |  |
| The cause of the DRP is related to the duration of treatment |  |  |
| C4.1 Duration of treatment too short | 4 (1.1) | - |
| C4.2 Duration of treatment too long | 40 (11) | 2 (5.1) |
| C8: Other: |  |  |
| C8.1 No or inappropriate outcome monitoring | - | 2 (5.1) |
| C8.2 Cost-effective alternatives available | - | 4 (12.8) |
| C8.3 No obvious cause | 2 (0.5) | - |
| **The Planned Interventions** |  |  |
| I3: At drug level |  |  |
| I3.1 Drug changed to | 102 (25.8) | - |
| I3.2 Dosage changed to | 55 (13.9) | - |
| I3.3 Formulation changed to | 4 (1) | - |
| I3.4 Instructions for use changed to | 85 (21.5) | - |
| I3.5 Drug paused or stopped | 86 (21.8) | - |
| I3.6 Drug started | 16 (4) | - |
| I4.1: Increased monitoring | 47 (11.9) | - |

**Table 3.** Severity of identified errors.

| Characteristic | Evaluation | Improvement |
| --- | --- | --- |
|  | N (%) | N (%) |
| **A. Potentially Lethal** |  |  |
| A1. High potential for life-threatening adverse effects/reactions | - | - |
| A2. Potentially lifesaving drug at a dosage too low for the disease being treated | 1 (0.3) | - |
| A3. High dosage (>10 times normal) of drug with low therapeutic index | - | - |
| **B. Serious** |  |  |
| B1. Route of administration could lead to severe toxicity | - | - |
| B2. Low dosage of drug for serious disease in patient with acute distress | - | - |
| B3. High dosage (4-10 times normal) of drug with low therapeutic index | - | - |
| B4. Dosage resulted in serum drug concentration in potentially toxic range | - | - |
| B5. Drug or combinations could exacerbate the patient's condition (related to warnings or contraindications) | 50 (13.7) | 1 (2.6) |
| B6. Misspelling or mix-up in medication order could lead to dispensing of wrong drug | - | - |
| B7. Documented allergy to drug | - | - |
| B8. High dosage (>10 times normal) of drug without low therapeutic index | 1 (0.3) | - |
| **C. Significant** |  |  |
| C1. High dosage (1.5-4 times normal) of drug with low therapeutic index | - | 1 (2.6) |
| C2. Drug dosage too low for patient's condition | 17 (4.7) | 3 (7.7) |
| C3. High dosage (1.5-10 times normal) of drug without low therapeutic index | 33 (9.1) | - |
| C4. Errant dual-drug therapy for single condition | 9 (2.5) |  |
| C5. Inappropriate dosage interval | 37 (10.2) | 1 (2.6) |
| C6. Omission from medication order | 15 (4.1) | 13 (33.3) |

**Table 3** (Continued)
| Characteristic | Evaluation | Improvement |
| --- | --- | --- |
|  | N (%) | N (%) |
| C7. Combination of drugs or drug might cause significant ADR to specific patient | 91 (25) | 11 (28.2) |
| D. Minor | - | - |
| D1. Incomplete information in medication order | - | - |
| D2. Unavailable or inappropriate dosage form | 2 (0.5) | - |
| D3. Non-formulary drug | 70 (19.2) | - |
| D4. Noncompliance with standard formulations and hospital policies | - | - |
| D5. Unnecessary instructions for specific time dosing | 12 (3.3) | 1 (2.6) |
| D6. Combination of drugs or drug might cause minor ADR to specific patient | 26 (7.1) | 2 (5.1) |
| E. No drug-related error |  |  |
| E1. Cost savings only | - | 4 (10.2) |
| E2. Increased monitoring required | - | 2 (5.1) |

<sup>a</sup> Adopted from Abdel-Qader (2010) and modified

**Table 4.** Characteristics of identified DRPs in service evaluation.

|  | N (%) |
| --- | --- |
| Number of patients according to identified DRPs |  |
| 0 | 307 (58) |
| 1 | 127 (24) |
| 2 | 62 (11.7) |
| 3 | 23 (4.4) |
| 4 | 6 (1.1) |
| 5 | 4 (0.8) |
| Drugs implicated with DRPs according to ATC main groups | 456 |
| A: Alimentary Tract and Metabolism | 86 |
| B Blood and blood forming organs | 32 |
| C: Cardiovascular System | 81 |
| D: Dermatologicals | 1 |
| G: Genito urinary system and sex hormones | 10 |
| H: Systemic hormonal preparations, excluding sex hormones and insulins | 9 |
| J: Anti-infective for systemic use | 37 |
| L: Antineoplastic and immunomodulating agents | 2 |
| M: Musculo-skeletal system | 18 |
| N: Nervous system | 118 |
| P: Antiparasitic products, insecticides and repellents | 0 |
| R: Respiratory system | 58 |
| S: Sensory organs | 4 |
| V: Various | 0 |
| Top 10 Drugs implicated with DRPs according to ATC 5th level |  |
| Bromazepam (N05BA08) | 19 |
| Esomeprazole (A02BC05) | 18 |
| Hydrochlorothiazide (C03AA03) | 14 |
| Clopidogrel (B01AC04) | 10 |
| Furosemide (C03CA01) | 10 |
| Gliclazide (A10BB09) | 9 |
| Glimepiride (A10BB12) | 9 |
| Olanzapine (N05AH03) | 9 |
| Alprazolam (N05BA12) | 9 |
| Zolpidem (N05CF02) | 8 |

### Interventions and Significance

In the evaluation project, 395 interventions were proposed by the pharmacist to a drug level (I3) and other intervention (I4), according to the PCNE classification. The vast majority were associated with drug level (I3, n=348, 88.1%) with only 11.9% of the total to include proposals for increased monitoring (Table 2). 202 of the total 364 DRPs were characterized assignificant (55.5%), 110 as minor (30.2%) and 51(14%) as serious. One potential error was defined as potentially lethal if not resolved.

### Improvement Project

The mean age of the four patients enrolled in the improvement project was 73 (59-84, SD=10.1). These patients have been taking a mean number of 6.5 drugs and we discovered 39 DRPs in their medication regimen (mean number of DRPs per patient= 9.75). “Adverse drug event possibly occurring” (P2.1, n=12, 30.8%) and “incomplete drug treatment in spite of existing indication” (C1.6, n=13, 33.3%) were the most recurrent among the problems and causes respectively. Through patient interviews we did not identify any causes of DRPs to be related with patient use, as they are defined in the PCNE classification system. Thus, we did not include this section in our classification neither in the first part, which would not be possible due to absence of information taken directly from patients, nor in the improvement part of this study. Almost one third (n=29, 74.36%) of assessed DRPs were assigned as significant and only one as serious (2.5%).

## DISCUSSION

In accordance with the reported data by Eurostat for Greece (2014), women that used prescribed medicines outweighed men in the service evaluation (Table 1). Moreover, the classification of all prescribed drugs (Table 1) was generally consistent with data on pharmaceutical consumption of prescribed drugs in Greece (2015), as they can be derived from OECD (2018). Two observed discrepancies in the service evaluation, when compared with the respective values provided by OECD (2018), included the low percentage of drugs of ATC-group B (blood and blood forming organs) and the high percentage of drugs of group R (respiratory system). The former evidence might be explained by the fact that low dose acetylsalicylic acid, a commonly used blood thinner, is available as an OTC product thus was not calculated in our prescribing data. For the latter, a contributor might be the mining region where the community pharmacy is located and its effect on prevalence of COPD and asthma (Sichletidis *et al.,* 2004). The 222 patients, with one or more DRPs, used more prescribed medicines (n=840) than the 307 patients in whom no DRP was identified (n=624) (Table 1). This aligns with studies that show that polypharmacy is associated with increased number of DRPs (Kovacevic *et al.,* 2017).

The overall population rate of encountered DRPs per patient (n=0.69) was comparable to two studies conducted at community pharmacies in Switzerland (n=0.68) (Eichenberger et al., 2010) and Sweden (n=0.48) (Montgomery *et al.,* 2008). However, other research projects conducted on similar settings showed a variety of identified DRPs per patient which ranged from 0.23 (Nicolas *et al.,* 2013) to 10.1 (Kwint *et al.,* 2012). Two other studies that took place at community pharmacies and used only medication records as data discovered 2.6 DRPs/patient (Krähenbühl *et al.,* 2008) and 4.1 DRPs/patient (Vinks *et al.,* 2009). Direct comparison of our results with the number of identified DRPs in other studies should be done with caution for a number of reasons. We observed differences in the study approach including variation in the definition of DRPs and also in the criteria utilized to identify actual or potential errors. In our study, we used a broad definition of DRPs and relied mostly on explicit criteria to increase the robustness of our results. The higher number of DRPs per patient detected in other studies might be explained by the fact that most of them used medical data and patients’ interviews as sources of information and a sample of polymedicated patients.

The most frequently identified DRPs were related to treatment safety and a potential adverse drug event (P2.1, n=226, 62.1%) (Table 2). The low number of problems related to medication effectiveness (P1, n=94, 25.8%) was anticipated since we did not have definitive outcomes as far as the efficacy of currently prescribed medicines in the first part of the study. A DRP was assigned as P1.2 “effect of drug treatment not optimal” when it was clear that an alternative available option was more beneficial for that patient considering his comorbidities and co-prescribed medicines. The most frequent among the causes documented was inappropriate combination of drugs (C1.4, n=75, 20.6%), which essentially were drug-drug interactions (Table 2). The high number of drug-drug interactions might be explained by the lack of an embedded software in prescriber’s electronic system at the time of writing the prescription. Thus, it is up to the physician’s discretion to search and consequently appraise the clinical significance of any drug-drug interactions. We identified many drugs to be inappropriately prescribed for specific patients (C1.2, n=72, 19.8%). Drugs with little or no evidence base or drugs with a high potential to cause a DRP considering patients comorbidities and age were assigned as C1.2. A great number of causes (C3, n=104, 28.57%) was related to the dose selection including high drug dose (C3.2, n= 40, 11%) and dosage regimen too frequent (C3.4, n= 28, 7.7%). These problems seem preventable through pharmacy counseling underpinning the role of community pharmacists in primary care as experts in medicine use. The latest versions of PCNE are not compatible with older ones which render the direct comparison of problem and cause results between studies problematic (PCNE, 2019).

Among the drugs implicated in DRPs, those intended for the nervous system showed the most incidents (n=118) (Table 4) and are in line with findings from a study conducted at French community pharmacies (Rhalimi *et al.,* 2018) (Table 4). Four out of ten drugs most frequently implicated with a DRP in our study belonged pharmacologically to benzodiazepines, hypnotics or atypical antipsychotics (Table 4). We observed that benzodiazepines and z-drugs were prescribed for indications of generalized anxiety disorder (GAD) and insomnia respectively even in repeat prescriptions for elderly patients, in contrast with guidelines and SPC which recommend time constraint on their use. All the aforementioned drug classes have been correlated with falls resulting in fractures and hospitalization while their cessation might lead to prevention of falls (Hill and Wee, 2012). Proton pump inhibitors (PPIs) belong to a class of medications that largely contributed to DRPs (n=38). These drugs are frequently prescribed in high doses and for long duration due to the common misconception that no ADRs are related with them. However, emerging evidence show that their misuse is related with increased infections, achlorhydria resulting in reduced absorption of minerals and vitamins, and increased incidents of fractures (Maes *et al.,* 2017).

The intervention at a drug level (I3) was the most frequent and specifically the three most frequently recurring were drug change (I3.1), drug paused or stopped (I3.5) and change of instructions (I3.4) (Table 2). Proposal to change a drug (I3.1, n=102, 25.8%) was associated with medicines that would potentially create an issue and an alternative drug or drug class might prevent the predicted problem. Intervention to pause or stop a drug (I3.5, n=86, 21.8%) was mostly related with medicines which were inappropriately prescribed for long duration or for medicines we judged that their effectiveness or safety was questionable. Proposals for change in drug use instructions (I3.4, n=85, 21.5%) were mainly associated with inappropriate dosing intervals for drugs like dihydropyridines, sulfonylureas or angiotensin-receptor-blockers (ARBs) that could unnecessarily increase patient’s pill burden or even adversely affect his treatment effectiveness or safety. From all the DRPs that were appraised regarding their clinical significance (Table 3), 202 (55.5%) were assigned as significant, 110 (30.2%) as minor and 51 (14%) as serious. One DRP (0.3%) was characterized as potentially lethal concerning the low prescribed dose of injectable epinephrine for treating an anaphylactic reaction.

### Improvement project

In the service improvement we identified a high number of DRPs (9.75 DRPs/patient). However, due to the small sample size we cannot draw robust conclusions or compare with the generated results from other studies. Errors arising from potential prescribing omissions or from absence of appropriate monitoring could not be discovered in the first part and this highlights the importance of clinical information obtained directly from patients. DRPs associated with patient’s use and behavior were not identified; however, we assume that this was due to the small sample of our service evaluation and the reluctance of patients to reveal potential inappropriate use of their medicines. Comprehensive medication reviews, as carried out in the improvement part, are most likely to benefit elderly patients on multiple conditions or altered pharmacodynamic/pharmacokinetic characteristics requiring adjustments in their pharmacotherapy and this should be taken into account in future relevant studies.

### Limitations of the study

The study approach we followed posed certain limitations. Access to patient’s medication record was limited on those possessed by the pharmacy. The identification, classification and assessment of significance for each DRP has been exclusively conducted by the pharmacist thus is considered subjective. Consistency of results would increase by employing more trained pharmacists and reaching in a consensus for the clinical criteria and estimation of clinical significance for each encountered DRP. Over the counter (OTC) medicines, which according to a study (Eickhoff *et al*., 2012) were correlated with 1 DRP per 5 OTC products dispensed, were not included in our service evaluation. Certain NSAIDs, like ibuprofen, may be either prescribed by the physician or directly purchased by the patient as OTC medicines. As their acquisition cost is low many patients opt to buy them without a prescription. Considering that more than one third of dispensed prescription medicines (n=506, 34.6%) (Table 1) in the service evaluation was intended for the cardiovascular system and that NSAIDs are known to be a risk factor for adverse cardiovascular events (Graham *et al.,* 2005) and impaired renal function (Hörl, 2010), we estimate that the actual number of DRPs in our population might be higher. In the main part of this study we did not explore if cost-effective alternative medicines were available. Errors associated with the logistics of the prescribing and dispensing process, like missing signatures, or patient related issues, like reduced adherence, were not included in the study either. The outcomes of proposed interventions were not assessed since the acceptance of interventions proposed to the physicians was beyond the scope of this study. Even though the majority of prescriptions filled in nowadays in Greece are electronic certain hand-written prescriptions, usually containing low-cost controlled drugs, are also executed and were not included in our study.

## CONCLUSIONS

The documentation of DRPs is considered a process indicator of pharmaceutical care (PCNE, 2018) and with our study we recorded a substantial number of DRPs of various causes in prescription medicines. Even though these DRPs were potential and may not be manifested eventually in patients, their presence renders medication regimen either unsafe or suboptimal while patients’ confidence in their care may be compromised. By incorporating a structured framework during reviews and extra sources of information the community pharmacists may identify issues that could not have been revealed before. The process of systematic identification of DRPs and consequent resolution with physician’s collaboration is an opportunity for community pharmacists to deviate from their traditional role in dispensing and by applying their knowledge be a part of the developing primary care in Greece. Future studies should focus on measuring additionally the financial and clinical impact of potential and actual DRPs.

## Data Availability

All data produced in the present work are contained in the manuscript.

## Supplementary Material

**Supplement 1:**
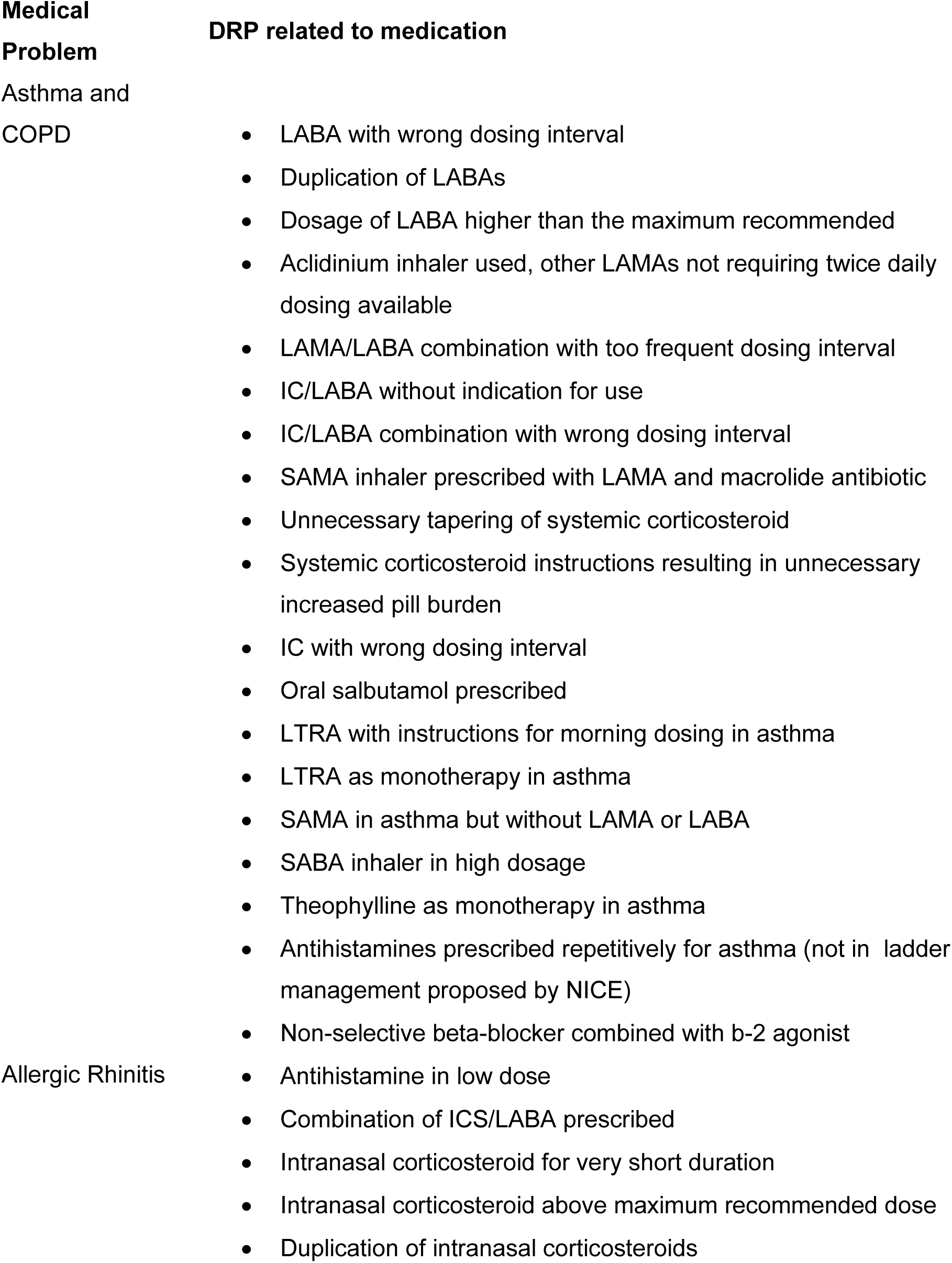
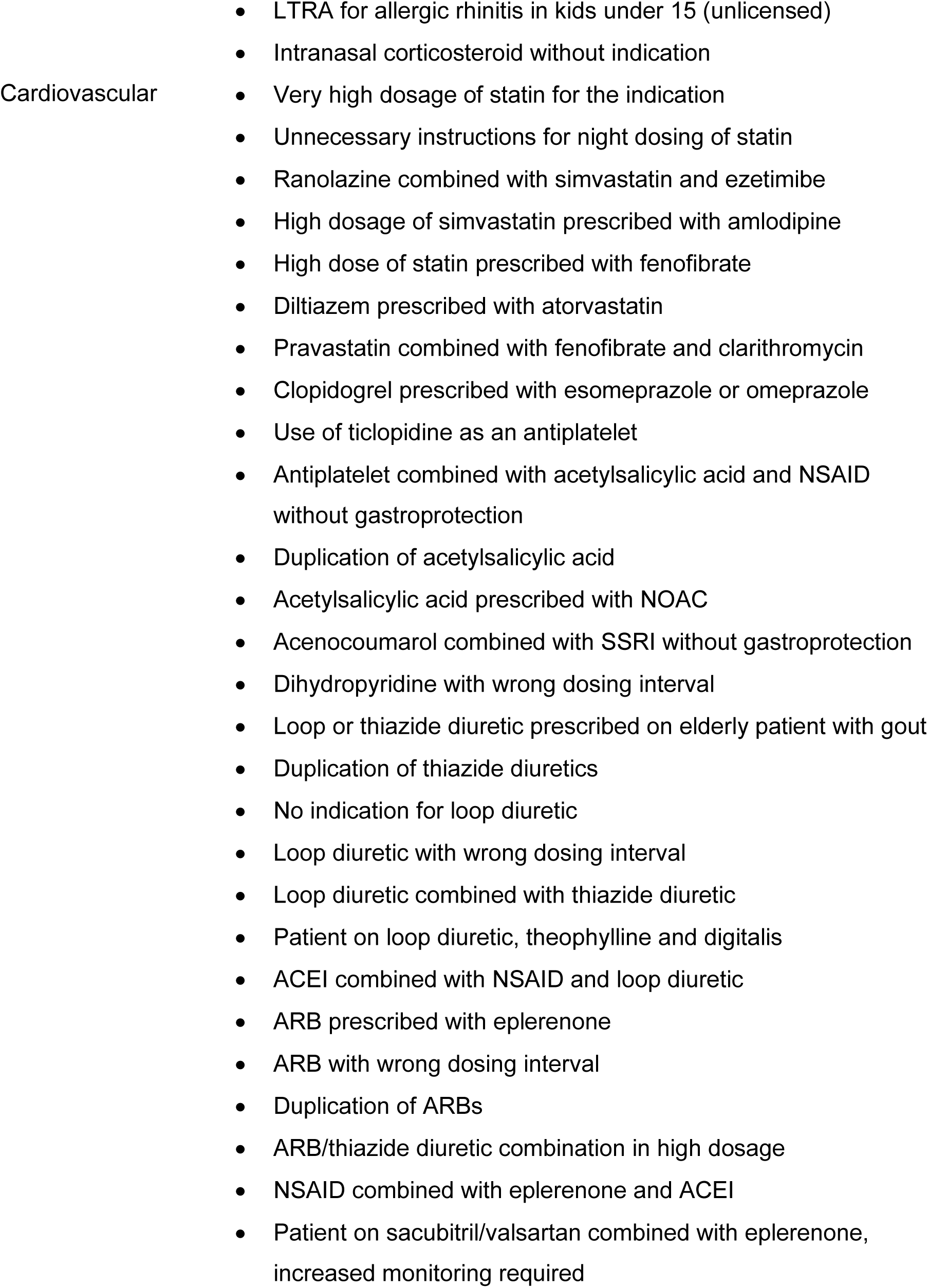
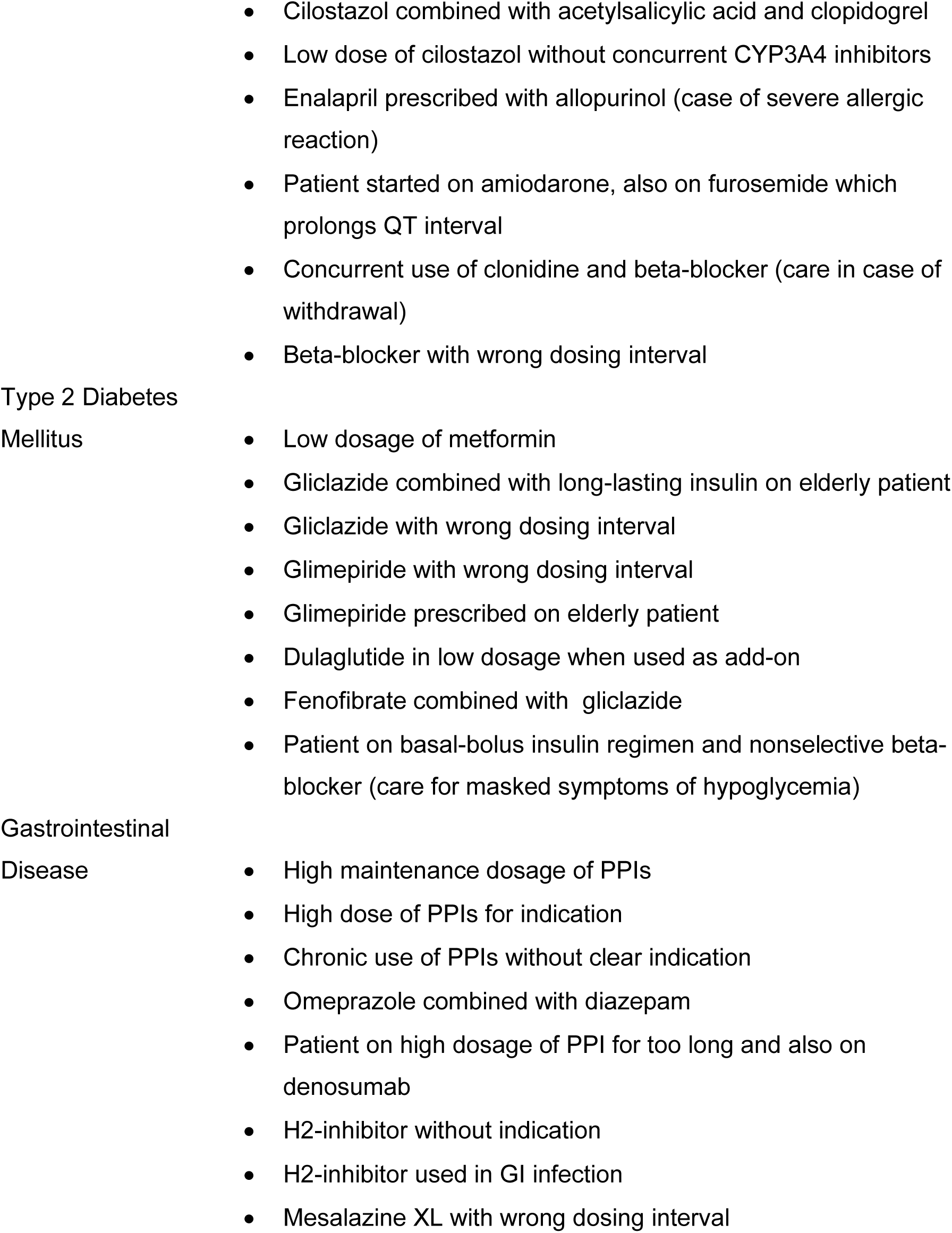
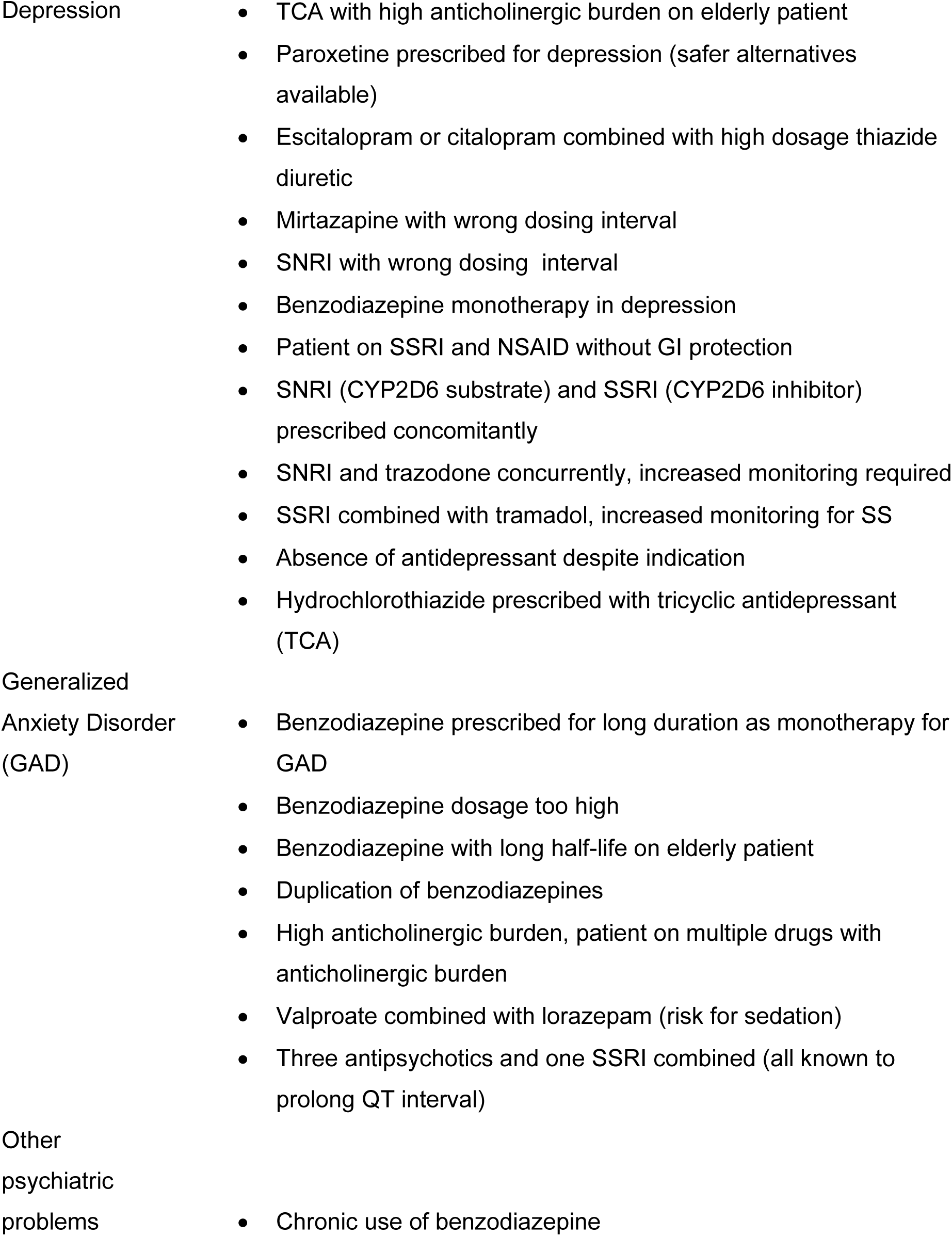
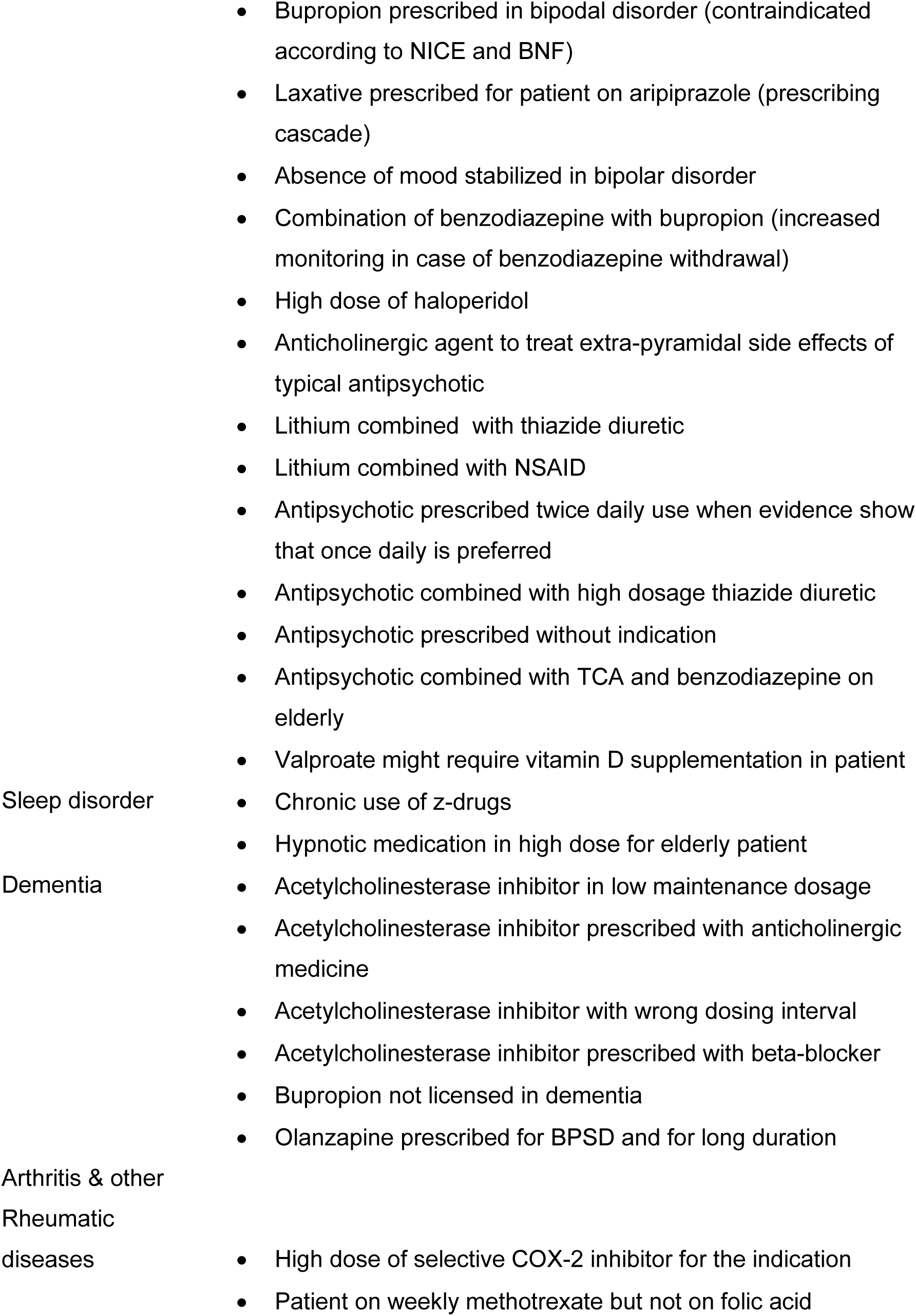
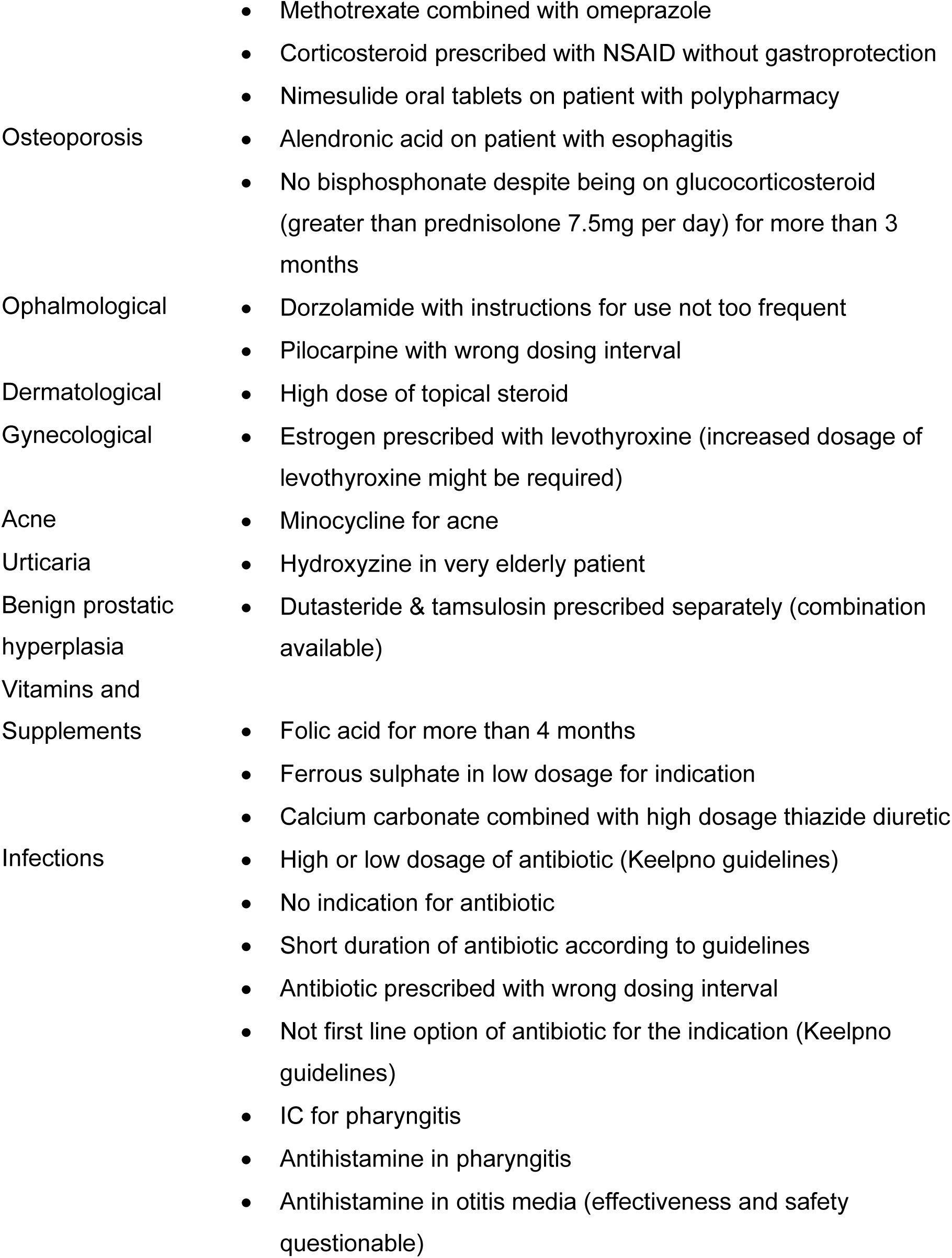
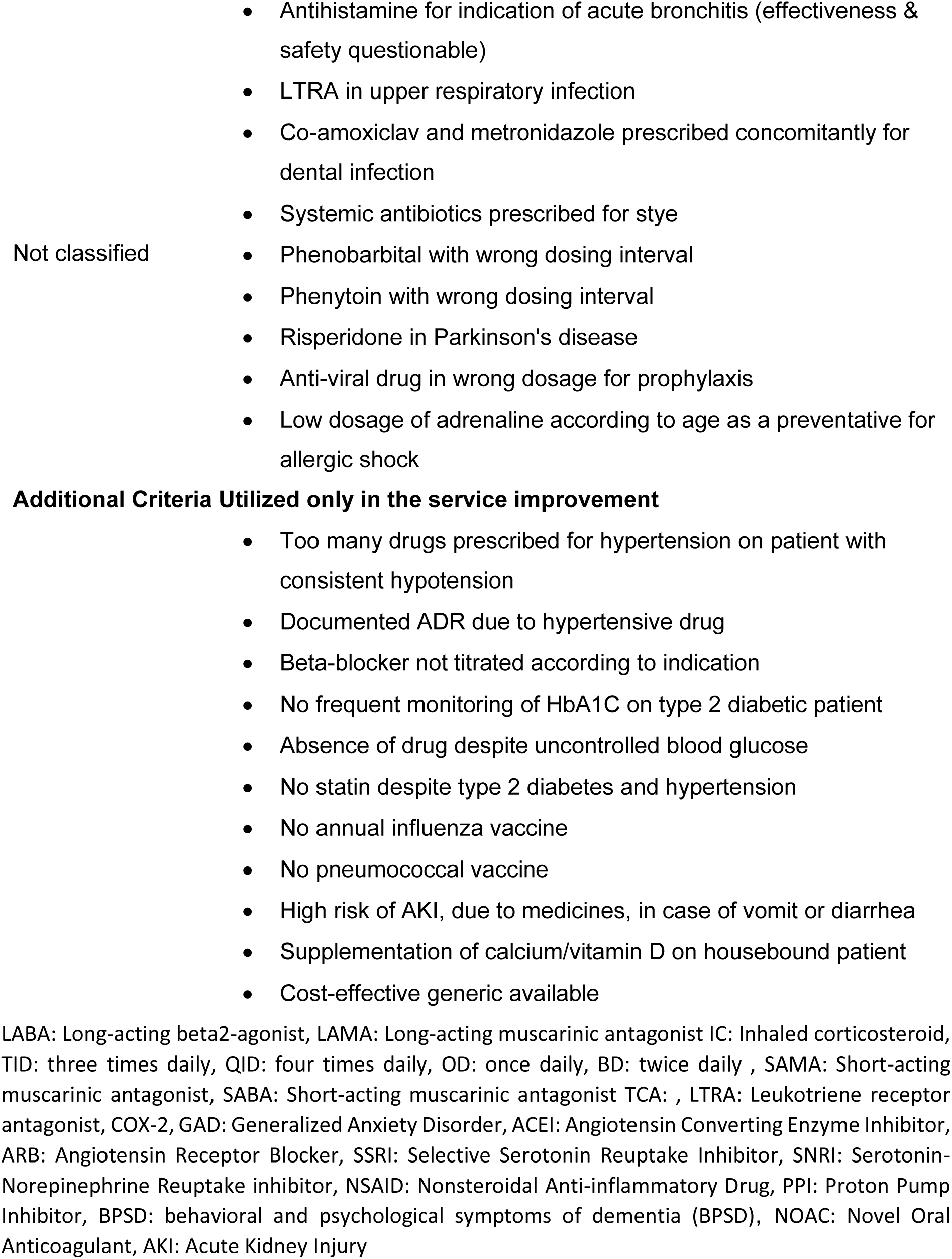
Clinical criteria utilized for potential DRPs in this study.

## List of References

1. Abdel-Qader, D. H., Harper, L., Cantrill, J.A. and Tully, M.P. (2010) ‘Pharmacists’ Interventions in Prescribing Errors at Hospital Discharge’, Drug Saf, 33(11) pp. 1027–1044 [online]. Available at: <https://link-springer-com.queens.ezp1.qub.ac.uk/article/10.2165%2F11538310-000000000-00000#Tab1> (Accessed: 24^th^ June 2019)

2. Aitken, M. and Gorokhovich (2012) ‘Advancing the responsible use of medicines: applying levels for change’ [online]. Available at: <https://papers.ssrn.com/sol3/papers.cfm?abstract_id=2222541>(Accessed: 29 April 2019)

3. Balourdos, D., Demertzis, N., Pierrakos, G., Kikilias, I. and Kosma, E. (2019) ‘Η χαμηλή γονιμότητα στην Ελλάδα, δημογραφική κρίση και πολιτικές ενίσχυσης της οικογένειας’. Greece [online]. Available at: <https://www.dianeosis.org/wp-content/uploads/2019/01/ekke_family_policies.pdf> (Accessed: 29 April 2019)

4. Barnett, K., Mercer, S.W., Norbury, M., Watt, G., Wyke, S., Guthrie, B. (2012) ‘Epidemiology of multimorbidity and implications for health care, research, and medical education: a cross-sectional study’, The Lancet, 380(9836), pp. 37–43 [online]. Available at:<https://www.thelancet.com/journals/lancet/article/PIIS0140-6736(12)60240-2/fulltext> (Accessed: 29 April 2019)

5. Barra, M., Scott, C.L., Scott, N.W., Johnston, M., Bruin, M., Nkansah, N., Bond, C.M., Matheson, C.I., Rackow, P. and Williams, A.J. (2018) ‘Pharmacist services for non-hospitalized patients’, Cochrane Database of Syst Rev [online]. Available at: <https://www.cochranelibrary.com/cdsr/doi/10.1002/14651858.CD013102/full?highlightAbstract=pharmacist%7Cwithdrawn%7Cservic%7Cservices%7Cserviz> (Accessed: 29 April 2019)

6. Basger B.J, Moles, R. J. and Chen T.F. (2014) ‘Application of drug-related problem (DRP) classification systems: a review of the literature’, Eur J of Clin Pharmacol 70(7), pp. 799–815, [online]. Available at: <https://link-springer-com.queens.ezp1.qub.ac.uk/article/10.1007%2Fs00228-014-1686-x> (Accessed: 24^th^ June 2019)

7. Baxter, K (ed) Stockley’s Drug Interactions, [online]. London: Pharmaceutical Press <www.medicinescomplete.com> (Accessed: 26th June 2019)

8. Cipolle, R. J., Strand, L. and Morley, P. (2012) Pharmaceutical Care Practice: The Patient-Centered Approach to Medication Management, 3rd edn., United States of America: The McGraw-Hill Companies

9. Donaldson, L.J., Kelley, E.T., Dhingra-Kumar, N., Kieny, M.P and Sheikh, A. (2017) ‘Medication without harm: WHO’s third global patient safety challenge’, The Lancet, 389, pp. 1680–1681, [online]. Available at: <https://www.thelancet.com/pdfs/journals/lancet/PIIS0140-6736(17)31047-4.pdf>(Accessed: 29 April 2019)

10. Eichenberger, P. M., Lampert, M. L., Kahmann, I. V., Foppe Van Mil, J. W. and Hersberger, K. E. (2010) ‘Classification of drug-related problems with new prescriptions using a modified PCNE classification system’, Pharm World Sci, 32(3), pp. 362–372, [online]. Available at:<https://link-springer-com.queens.ezp1.qub.ac.uk/article/10.1007%2Fs11096-010-9377-x> (Accessed: 24th June 2019)

11. Eickhoff, C., Hämmerlein, A., Griese, N. and Schulz, M. (2012) ‘Nature and frequency of drug-related problems in self-medication (over-the-counter drugs) in daily community pharmacy practice in Germany’ *Pharmacoepidemiology and Drug Saf*, 21, pp. 254–260 [online]. Available at: <https://link-springer-com.queens.ezp1.qub.ac.uk/article/10.1007%2Fs11096-013-9769-9> (Accessed: 24^th^ June 2019)

12. Economou, C., Kaitelidou, D., Karanikolos, M. and Maresso, A (2017) ‘Greece: Health system review’ *Health Systems in Transition* 19(5), pp. 1–192, [online]. Available at:<http://www.euro.who.int/data/assets/pdf_file/0006/373695/hit-greece-eng.pdf>(Accessed: 29 April 2019)

13. EMC (2019) Summary of Product Characteristics, Available at: <https://www.medicines.org.uk/emc> (Accessed: 26th June 2019)

14. European Commission (2015) *Demography Report* – Luxemburg: European Commission [online]. Available at: <https://ec.europa.eu/eurostat/documents/3217494/6917833/KE-BM-15-003-EN-N.pdf/76dac490-9176-47bc-80d9-029e1d967af6>(Accessed: 29 April 2019)

15. Eurostat (2014) *Medicine use statistics*, Available at: <https://ec.europa.eu/eurostat/statistics-explained/index.php?title=Medicine_use_statistics> (Accessed: 24^th^ June 2019)

16. Eurostat (2019) *Population (Demography, migration and projections)*, Available at: <https://ec.europa.eu/eurostat/web/population-demography-migration-projections/visualisations> (Accessed: 26^th^ June 2019)

17. Garfield, S., Reynolds, M., Dermont, L. and Franklin, B. D. (2013) ‘Measuring the severity of prescribing errors: a systematic review’ *Drug Saf*, 36, pp. 1151-1157 [online]. Available at:<https://www.ncbi.nlm.nih.gov/pmc/articles/PMC3834169/> (Accessed: 25^th^ June 2019)

18. Graham, D. J., Campen, D., Hui, R., Spence, M., Cheetham, C., Levy, G., Shoor, S. and Ray, W. (2005) ‘Risk of acute myocardial infarction and sudden cardiac death in patients treated with cyclo-oxygenase 2 selective and non-selective non-steroidal anti-inflammatory drugs: nested case-control study”, The Lancet, 365(9458), pp. 475–481 [online]. Available at:<https://www.sciencedirect.com/science/article/pii/S0140673605178647> (Accessed: 25^th^ June 2019)

19. Hamilton, H., Gallagher, P. and Ryan, C. (2011) ‘Potentially inappropriate medications defined by STOPP criteria and the risk of adverse drug events in older hospitalized patients’ Arch Intern Med 171 (11), pp. 1013–1019, [online]. Available at:<https://jamanetwork.com/journals/jamainternalmedicine/article-abstract/227481> (Accessed: 24^th^ June 2019)

20. Hill, K. D. and Wee, R. (2012) ‘Psychotropic drug-induced falls in older people: a review of interventions aimed at reducing the problem’, Drugs Aging 29(1), pp. 15–30 [online]. Available at: <https://www.ncbi.nlm.nih.gov/pubmed/22191720> (Accessed: 24^th^ June 2019)

21. Hörl (2010) ‘Nonsteroidal anti-inflammatory drugs and the kidney’ Pharmaceuticals (Basel), 3(7), pp. 2291–2321 [online]. Available at:<https://www.ncbi.nlm.nih.gov/pmc/articles/PMC4036662/> (Accessed: 25^th^ June 2019)

22. Joint Formulary Committee. British National Formulary (online) London: BMJ Group and Pharmaceutical Press <http://www.medicinescomplete.com> (Accessed at 26th June 2019)

23. KEELPNO (2015) Guidelines for the diagnosis and treatment of infectious disease- Greece, Athens: Greek Society for Infections [online]. Available at:<http://www.thriassio-hosp.gr/wp-content/uploads/2018/03/%CE%9A%CE%95%CE%95%CE%9B%CE%A0%CE%9D%CE%9F-%CE%9A%CE%9F-2015.pdf> (Accessed: 25^th^ June 2019)

24. Kovacevic, S.V, Milijkovic, B., Kovacevic, M., Golubovic, B., Jovanovic, M., Vucicevic, K. and Gier, J.J. (2017) ‘Evaluation of drug-related problems in older polypharmacy primary care patients’, J Eval Clin Pract, 23(4), pp. 860–865 [online]. Available at:<https://www.ncbi.nlm.nih.gov/pubmed/28370742> (Accessed: 26^th^ June 2019)

25. Krähenbühl, J. M., Decollogny, A. and Bugnon, O. (2008) ‘Using the costs of drug therapy to screen patients for a community pharmacy-based medication review program’, Pharm World Sci, 30, pp.816–822 [online]. Available at: <http://link.springer.com/10.1007/s11096-008-9232-5> (Accessed: 24^th^ June 2019)

26. Laliotis, I., Ioannidis, J. P. A., Stavropoulou, C. (2016) ‘Total and cause-specific mortality before and after the onset of the Greek economic crisis: an interrupted time-series analysis’, The Lancet Public Health, 1(2), pp. e56–e65 [online]. Available at: <https://www.sciencedirect.com/science/article/pii/S2468266716300184> (Accessed: 29 April 2019)

27. Maes, M. L., Fixen, D. R. and Linnebur S. A. (2017) ‘Adverse effects of proton-pump inhibitor use in older adults: a review of the evidence’, Therapeutic Advances in Drug Safety 8(9), pp. 273–297 [online]. Available at:<https://www.ncbi.nlm.nih.gov/pmc/articles/PMC5557164/> (Accessed: 24^th^ June 2019)

28. Montgomery, A. T., Sporrong, S. K., Tully, M. P. and Lindblad, A. K. (2008) ‘Follow-up of patients receiving a pharmaceutical care service in Sweden’, J Clin Pharm Ther 33, pp. 653–662 [online]. Available at:<https://onlinelibrary-wiley-com.queens.ezp1.qub.ac.uk/doi/full/10.1111/j.1365-2710.2008.00965.x> (Accessed: 24^th^ June 2019)

29. Nicolas, A., Eickhoff, C., Griese, N. and Schulz, M. (2013) ‘Drug-related problems in prescribed medicines in Germany at the time of dispensing”, Int J Clin Pharm 35, pp. 476–482 [online]. Available at:<https://link-springer-com.queens.ezp1.qub.ac.uk/article/10.1007%2Fs11096-013-9769-9> (Accessed: 24^th^ June 2019)

30. Norwegian Institute of Public Health (2017) *ATC/ DDD Index* 2018 Available at: <https://www.whocc.no/atc_ddd_index/>

31. OECD (2018) *Pharmaceutical market: pharmaceutical consumption*, Available at: <https://stats.oecd.org/Index.aspx?DataSetCode=HEALTH_PHMC> (Accessed: 24^th^ June 2019)

32. OECD/European Observatory on Health Systems and Policies (2017) *Greece: Country Health Profile 2017, State of Health in the EU -* Paris: OECD Publishing/Brussels: Observatory on Health Systems and Policies [online]. Available at:<https://ec.europa.eu/health/sites/health/files/state/docs/chp_gr_english.pdf> (Accessed: 25^th^ June 2019)

33. O’Mahony, D., O’Sullivan, D., Byrne, S., O’Connor, M. N., Ryan, C. and Gallagher, P. (2015) ‘STOPP/START criteria for potentially inappropriate prescribing in older people: version 2’ *Age Ageing*, 44(2), pp. 213–218, [online]. Available at:<https://www.ncbi.nlm.nih.gov/pmc/articles/PMC4339726/> (Accessed: 24^th^ June 2019)

34. Pharmaceutical Care Network Europe (2019) *The PCNE Classification V 8.03* [online]. Available at:<https://www.pcne.org/upload/files/318_PCNE_classification_V8-03.pdf>(Accessed: 29 April 2019)

35. Rhalimi, M., Rauss, A. and Housieaux, E. (2018) ‘Drug-related problems identified during geriatric medication review in the community pharmacy’, Int J Clin Pharm 40, pp.109–118 [online]. Available at:<https://link.springer.com/article/10.1007%2Fs11096-017-0571-y> (Accessed: 24th June 2019)

36. Ryan, C., O’Mahony, D., O’Donovan, D. O., O’Grady, E., Weedle, P., Kennedy, J. and Byrne S. (2013) ‘A comparison of the application of STOPP/START to patients’ drugs lists with and without clinical information’, Int J Clin Pharm 35, pp. 230–235 [online]. Available at:<https://link-springer-com.queens.ezp1.qub.ac.uk/article/10.1007%2Fs11096-012-9733-0> (Accessed: 25th June 2019)

37. Scottish Government Polypharmacy Model of Care Group. Polypharmacy Guidance, Realistic Prescribing 3 rd Edition, 2018. Scottish Government [online]. Available at: <https://www.therapeutics.scot.nhs.uk/wp-content/uploads/2018/09/Polypharmacy-Guidance-2018.pdf>(Accessed: 25th June 2019)

38. Sichletidis, L., Tsiotsios, I., Chloros, D., Daskalopoulou, E., Ziomas, I., Michailidis, K., Kottakis, T.H. and Palladas, P. (2004) ‘The effect of environmental pollution on the respiratory system of lignite miners: a diachronic study.’ Med Lav, 95(6), pp. 452–464 [online]. Available at:<https://www.ncbi.nlm.nih.gov/pubmed/15736306> (Accessed: 30^th^ June 2019)

39. SFEE (2017) The Pharmaceutical Market in Greece: facts and figures *-* Greece [online]. Available at:<https://www.sfee.gr/wp-content/uploads/2018/03/FF2017-eng-1.pdf> (Accessed: 25^th^ June 2019)

40. Taché, S. V., Sönnichsen, A. and Ashcroft, D. M. (2011) ‘Prevalence of adverse drug events in ambulatory care: a systematic review’ *Ann of Pharmacother*, 45(7-8) [online]. Available at:<https://journals.sagepub.com/doi/abs/10.1345/aph.1p627>(Accessed: 29 April 2019)

41. Vinks, T. H. A. M., Egberts, T. C. G., Lange, T. M. and Konig, F. H. P. (2009) ‘Pharmacist-based medication review reduces potential drug-related problems in the elderly’ *Drugs Aging*, 26(2), pp. 123–133 [online]. Available at:<https://link-springer-com.queens.ezp1.qub.ac.uk/article/10.2165%2F0002512-200926020-00004> (Accessed: 24^th^ June 2019)

